# Efficacy and Moderators of Mindfulness-Based Cognitive Therapy (MBCT) in ‘Difficult to Treat’ Depression: Protocol for a Systematic Review and Individual Patient Data Meta-Analysis of Randomised Controlled Trials

**DOI:** 10.1101/2025.06.09.25329249

**Authors:** Thorsten Barnhofer, Maria Niemi, Johannes Michalak, Maria Velana, J. Mark G. Williams, Alberto Chiesa, Stuart Eisendrath, Kevin Delucchi, Zindel V. Segal, Mira Cladder-Micus, Anne Speckens, Ali Akbar Foroughi, Mauro Garcia-Toro, Jesus Montero-Marin, Barnaby D. Dunn, Clara Strauss, Florian A. Ruths, Mary Ryan, Mathias Harrer

## Abstract

**Introduction:** About 30% of depressed patients suffer from a protracted course in which the disorder continues to cause significant burden despite treatment efforts. While originally developed for relapse prevention, Mindfulness-Based Cognitive Therapy (MBCT) has increasingly been investigated in depressed patients with such ‘difficult-to-treat’ courses. This is a protocol for an individual participant data (IPD) meta-analysis aiming to determine efficacy and potential moderators of MBCT treatment effects in this group based on evidence from randomised controlled trials.

**Methods and analysis:** Systematic searches in PubMed, Web of Science, Scopus, PsycINFO, EMBASE, and the Cochrane Controlled Trials Register for randomised controlled trials were completed on 17 June 2024. Authors of identified studies have contributed IPD and data extractions have been completed. An update search will be conducted immediately before the start of data analyses. We will investigate the following outcomes: (a) self-reported and observer-reported severity of depression symptomatology, (b) remission, (c) clinically meaningful improvement and deterioration. One-stage and two-stage IPD meta-analyses will be conducted with one-stage models using the observed IPD from all studies simultaneously as the primary approach. One-stage IPD models will include stratified study intercepts and error terms as well as random effects to capture between-study heterogeneity. Moderator analyses will test treatment-covariate interactions for both individual patient level and study level characteristics.

**Ethics and dissemination:** The results will inform understanding of the use of MBCT in patients with current ‘difficult-to-treat’ depression and will contribute to arguments in favour of or against implementing MBCT as a treatment for this group. They will be published in a peer reviewed journal and made available to stakeholders in accessible formats. No local ethical review was necessary following consultation with the Ethics and Governance Board of the University of Surrey. Guidance on patient data storage and management will be adhered to throughout.

**PROSPERO registration number:** CRD42022332039

**STRENGTHS AND LIMITATIONS OF THIS STUDY:**

- The chosen approach using IPD has several advantages over conventional meta-analyses including the possibility to assess whether individual participant level characteristics influence treatment effects with more specificity and power
- Limitations of the approach include the fact that inconsistencies across studies regarding the format and type of variables reported may restrict our ability to investigate moderators of treatment effects
- Transformation of outcome measures to a common metric across all studies may come with a small loss of information.

## INTRODUCTION

Major Depressive Disorder (MDD) is highly prevalent, debilitating [1], and in about 30% of patients takes a recurrent or chronic course in which established treatments fail to bring sustained remission [2]. Recently framed under the broader heuristic of difficult-to-treat depression (DTD) [3, 4], such courses are associated with continuing functional impairment, reduced quality of life, and significantly increased risk for chronic physical and neurodegenerative disorders [5, 6].

While treatment approaches for DTD have traditionally been dominated by pharmacological strategies [7], it is now increasingly acknowledged that effective management requires a broader emphasis on working with symptoms, maximising function and minimizing burden [4]. Psychological interventions have an important contribution to make in this context [8] and it has been suggested that Mindfulness-Based Cognitive Therapy (MBCT [9]), an eight-week treatment that combines elements of cognitive therapy with mindfulness meditation, may be particularly suited for this purpose. Using a mental training approach, the intervention aims to provide patients with portable skills that enable them to disengage from habitual maladaptive responses and respond more adaptively to negative mood and stress.

MBCT was originally introduced for the prevention of relapse in patients who are currently in remission and has been proven to be effective for this purpose [10], particularly in patients with highly recurrent courses (a use that is endorsed by treatment guidelines internationally). Early trials restricted the intervention to patients who were currently in full remission as it was assumed that intensive mindfulness practice may be too demanding for those who are suffering from current symptoms [11, 12]. However, as patients who are at high risk of relapse are likely to show residual symptoms between episodes, this restriction was later loosened, and analyses across studies for relapse prevention show that treatment effects increase, rather than decrease, with level of residual symptoms [10]. From a theoretical perspective, this observation is in line with the view that mindfulness skills serve to buffer the effects of negative mood and stress [13, 14], which implies that effects should become more visible under conditions of higher symptoms or stress.

Based on this reasoning, research has extended the use of MBCT to patients with current depression that has proven ‘difficult-to-treat’. Preliminary studies investigating MBCT in treatment-resistant and chronic depression were published in 2008 [15] and 2009 [16] and several definitive randomised controlled trials have been conducted since then [17–22], suggesting that evidence has accumulated to a point where the use of MBCT as a treatment for patients with DTD can now be considered for guideline endorsement and wider implementation. It seems timely therefore to bring data from existing randomised controlled trials together and analyse outcome across studies.

For this purpose, we have pooled individual patient data (IPD) from these studies to conduct meta-analyses with one-stage random effects meta-analysis of independent patient data (IPD) as our primary approach given its advantages over conventional aggregate data meta-analyses [23–26]. The main aim of the analyses is to establish efficacy of MBCT for patients with DTD as compared to the control conditions utilised in the existing trials. Given the need for both effective symptom reduction and the maintenance of gains, we will aim to determine immediate effects at the end of the eight-week MBCT intervention (post-treatment) and effects in the longer-term after patients have finished the intervention and are free to continue engaging in mindfulness practice by themselves (follow-up).

An important advantage of IPD meta-analysis is that it allows to examine moderator or subgroup questions across the pooled sample and can thus serve to overcome limitations in sample size and power for answering these questions in the original studies [27]. Information about potential moderators of treatment effects, such as severity of depression at entry to treatment, is important as it can potentially help with implementation by providing more precise information about relative indications or contraindications of the intervention. A further aim of this IPD meta-analysis is therefore to explore effects of potential individual patient level (e.g. sociodemographic and clinical characteristics) and study level (e.g. delivery format) moderators of treatment effects in the pooled data set.

As the clinical concept of DTD does not link to an objective way of assessment, the current meta-analysis will include studies that have used a range of conventional definitions falling under the wider concept of DTD, including selecting participants based on treatment non-response/remission, treatment resistance or the presence of chronic depression. Subgroup analyses will allow us to determine the extent to which effects found in the aggregated data generalise across these conventional categories.

## METHODS

### General approach

This IPD meta-analysis is registered on PROSPERO (registration number: CRD42022332039; date of registration in PROSPERO: 24 May 2022) and any key changes or amendments will be documented there. The protocol is registered retrospectively as searches to identify eligible papers were already conducted in 2024, with the last search having been performed on 17 June 2024 and an update search planned immediately before the start of data analysis. Corresponding authors of the selected studies were contacted and asked to provide raw data from their studies. The Preferred Reporting Items for Systematic Reviews and Meta-Analyses IPD statement [28] will be followed for the reporting of this study.

### Eligibility criteria

#### Types of studies to be included

This IPD meta-analysis will include only peer-reviewed RCTs, published in English.

There will be no restriction on the type of recruitment settings.

#### Type of intervention

Included studies will have investigated the effects of manualised mindfulness-based cognitive therapy (MBCT) alone or as an adjunct treatment to treatment-as-usual (TAU) [29]. MBCT is a psychological group-based intervention that combines intensive training in mindfulness meditation with elements of cognitive behavioural therapy (CBT) for depression. Mindfulness can be operationally defined as non-judgemental awareness of present moment experience [30]. As MBCT had been originally designed to prevent relapse into depression and not to reduce symptoms during an episode, we will accept studies that introduce minor changes to the manual (e.g. incorporating specific support for managing current depressive symptoms), as well as standard manualised MBCT, but not other types of mindfulness-based interventions, and include MBCT studies regardless of delivery format (e.g. physical group setting versus videoconference).

#### Types of comparators

This may include TAU for depression, including pharmacological and psychological treatments (e.g., CBT) or active controls. The latter will include established interventions as well as control interventions designed to mimic effects of mindfulness-based interventions in some or all aspects with the exception of the core element of mindfulness.

#### Participants

We will include studies that recruited adults with current Major Depressive Disorder (MDD) [31, 32] and studies which included mixed groups of patients with residual depressive symptoms or full episodes in the context of a chronic or treatment-resistant course of the disorder. In the latter case, participants with only residual symptoms will be excluded from the primary analysis approach (one-stage meta-analysis). As DTD does not link to an objective way of assessment, we will opt for a maximally inclusive interpretation of the term and include any studies that selected patients based on previous treatment non-response/remission, treatment-resistance or chronicity. We will exclude studies with participants below the age of 18 and studies that have exclusively recruited participants over the age of 65 given that the factors involved in the onset, maintenance and recurrence of depression in these age groups can differ from those in adults within this age bracket [33, 34].

### Outcomes

The main type of outcome will be severity of depressive symptomatology as assessed using standard depression outcome measures. Standard depression outcome measures may include self-report measures and observer-rated measures. If both types of measures are available, we will investigate effects on both types of outcome measures. If several measures are available for a single study, preference will be given to the measure that was used as the primary outcome of the original study. To compare outcomes across studies using different outcome measures, we will transform scores from these measures into standardised scores using the common metric developed by Wahl et al. [35]. Where a measure is not part of the common metric approach, we will transform the measure into one that is part of the common metric using published conversion tables [36], and then derive the common metric.

To facilitate clinical interpretation of results, we will derive dichotomous outcomes reflecting remission, improvement and deterioration. To apply a common rule for all studies, common metric scores will be converted to the PHQ-9 and dichotomous outcomes computed based on this measure. Following conventions for the PHQ-9 used in pragmatic trials and health services, remission will be defined as a score of less than 10 [37, 38], i.e. a shift from “caseness” to a non-case score. Definition of clinically meaningful change will follow suggestions from recent trials in treatment-resistant depression, which has indicated a threshold of 6 or more points [39, 40]. We will therefore define clinically meaningful improvement as a reduction of 6 or more points and clinically meaningful deterioration as an increase of 6 or more points on the PHQ-9.

Secondary outcomes such as mindfulness, self-compassion and quality of life will be collected and may be used for future analysis.

### Moderators

We will also investigate potential moderators of treatment response, i.e. factors that determine whether a person responds better to MBCT as compared to the control condition or vice versa. These will include potential moderators on the individual patient level (e.g. sociodemographic characteristics, history of childhood trauma, and clinical characteristics) and on the study level. We will include patient level characteristics in the analyses, if they are reported across a sufficient number of studies and datasets (see statistical analysis section for criteria).

Sociodemographic and clinical characteristics that we aim to investigate include age [41], gender [42], marital status [43], education [44], employment [45], childhood adversity [46], age at onset of depression [47], number of previous episodes [48], duration of current episode [49], current suicidality [50], current antidepressants [51], past antidepressants and treatment-resistance to antidepressants [52], co-morbid anxiety disorder, comorbid obsessive-compulsive disorder, co-morbid post-traumatic stress disorder, co-morbid substance abuse or dependence, co-morbid eating disorder, co-morbid personality disorder [41], and level of depressive symptoms at entry [53]. Study level characteristics will include type of delivery (remote versus face-to-face), type of control condition (TAU, active control), type of DTD (non-response/remission, treatment-resistance, chronic depression) and study quality.

### Timing of outcome assessments

As follow-up periods of the studies differ, we will group follow-up periods in the following categories: 8 to 10 weeks post-randomisation (post-treatment), 11 to 34 weeks post-randomisation (follow-up 1), and 35 to 60 weeks post-randomisation (follow-up 2).

### Searches and study selection

We searched PubMed, Web of Science, Scopus, PsychINFO, EMBASE, and the Cochrane Controlled Trials Register using index and free terms, jointly with Boolean operators, on four tiers, namely: (1) depressive disorder, (2) non-response, treatment resistance and chronicity, (3) mindfulness-based cognitive therapy, and (4) randomised-controlled trial. The search terms used for the different databases are shown in the appendix.

We searched for any study published up until the day of the search, on 17 June 2024. There was no lower time limit. Initial screening of titles was performed by two independent reviewers (TB, JM), with the publication abstract being obtained if selected by at least one reviewer. Abstracts were screened by the same two independent reviewers, with disagreements about eligibility being referred to a third reviewer whose decision was final. An update search will be conducted immediately before the start of data analysis.

### Risk of bias assessment

The validity of the studies will be assessed using the Risk of Bias tool by the Cochrane Collaboration (version 2) [54, 55]. Possible sources of bias assessed by the tool are (1) random sequence generation, (2) allocation concealment, (3) blinding of outcome assessors, (4) incomplete outcome data, (5) selective outcome reporting, and (6) other threats to validity including lack of compliance and different timing of outcome assessments. Data from the published papers of the trials will be evaluated by two independent assessors.

Agreement rate will be reported, and any disagreements will be solved through discussion. A sensitivity analysis will be performed excluding studies judged to be at high risk of bias, or where risk of bias is unclear.

### Patient and public involvement

A lived experience advisory group, led by MR and consisting of experts who have been involved in previous trials of MBCT, will be consulted for the interpretation of study results and their dissemination. A coproduced lay summary of results will be written for dissemination alongside papers for peer review journals.

### IPD data collection and aggregation

All corresponding authors of the articles selected for inclusion were contacted via email by TB with an invitation to participate in the review and to share the IPD from their primary study. The standard letter described all relevant details of the study including its purpose, main research questions, and the variables of interest and ended with a request to share the raw data from the study. Corresponding authors of studies were invited to serve as collaborators for the project. If corresponding authors did not respond within four weeks, we sent a second email request. If this had failed, a second author was contacted. Apart from one study, whose authors could not be contacted, none of the selected studies required contact attempts beyond a second author and the responding authors agreed to share their data.

### Data checking and integrity

Authors were requested to provide a codebook or data dictionary explaining variable names, coding of values, measurement units etc. After accepting the invitation to collaborate and signing the data transfer agreement, the study authors shared their IPD via secure data transfer portal at the University of Surrey. Data is stored securely on University of Surrey servers in accordance with the European Union’s General Data Protection Regulation Act and the UK’s Data Protection Act 2018.

Received datasets will be reviewed to assess completeness and checked against the reported data from relevant study publications. The numbers of participants, overall and by arm, will be confirmed to correspond with the numbers in the study publication. Ranges of values will be inspected for clearly erroneous data. Baseline participant demographic data will be inspected by arm, with the aim of checking that the IPD matches the data reported in the publication, in terms of the numbers of participants with reported data, and overall aggregated values for each variable. Discrepancies will be referred to the study authors for investigation and any remaining discrepancies will be recorded.

Although the main search and initial receipt of IPD occurred before the submission of this protocol for publication, the protocol was registered in PROSPERO (CRD42022332039) beforehand. No data analysis or outcome-driven decisions were made prior to finalizing the protocol, and the methods described here were pre-specified prior to data inspection.

### Data harmonization

Once datasets from the individual studies have been checked and outcomes standardised, they will be merged into the final dataset. The final dataset for analysis will be checked again for accuracy by a researcher in the study team.

## STATISTICAL ANALYSIS

To finalise the selection of moderator variables, we will determine presence of these variables across the individual data sets and decide on ways to standardise these variables where needed, for example by collapsing categories. Potential moderator variables will be included in the analyses if the variable has at least 40% available data in the final dataset and has been recorded by at least 4 studies.

To investigate the efficacy of MBCT compared to control conditions and test moderating effects of individual patient level and study level characteristics, we will conduct one-stage and two-stage IPD meta-analyses (IPD-MAs). Random-effects models will be used throughout because we expect high statistical heterogeneity due to variation in control groups (treatment as usual, active controls) and study populations (treatment non-responders/remitters, treatment-resistance, chronic depression).

The main research question of the study (whether MBCT is efficacious compared to control conditions) will be addressed via a maximum of three pairwise comparisons: (1) MBCT versus treatment-as-usual controls, (2) MBCT versus active controls, and (3) MBCT versus all controls.

By default, all analyses will be conducted in a Bayesian framework using JAGS (Just Another Gibbs Sampler)/BUGS (Bayesian analysis Using Gibbs Sampling) [56]. We chose a Bayesian approach because it offers greater flexibility in the parametrization of the (one-stage) IPD-MA models, and because this allows for a more intuitive interpretation of results based on posterior tail probabilities [57]. Across all analyses, findings will be considered “significant” when the credibility interval of the relevant effect does not include zero.

### Two-stage random effects meta-analysis

We will first conduct a conventional meta-analysis using data extracted from the published papers. We will calculate Hedges’ g to determine the effect sizes of the difference between the intervention and the control conditions reported in the papers. To investigate heterogeneity of effect sizes, we will calculate I^2^, the percentage of variation not attributable to sampling error, as well as 95% prediction intervals (PIs) around the pooled effect [58, 59]. Leave-one-out analyses will be conducted to identify potential outliers and/or influential studies. Small-study effects will be investigated visually using funnel plot inspection and the Egger’s test for asymmetry. A weakly informative Half-Cauchy (0,0.5) prior will be used for the between-study heterogeneity variance τ^2^ [60], and flat normal priors for all other parameters. A detailed justification for this prior setup has been derived in previous work [61].

### One-stage random effects meta-analysis using IPD

One-stage IPD meta-analysis models will be used as the primary analytic approach. To analyse the effects on common metrics-converted symptom severity scores, an identity-link linear mixed effects model with stratified intercepts and trial-specific (heteroscedastic) error terms will be used. Let *y_ik_* be the depressive symptom severity of patient *i* in trial *k* (with *k*=1,…, *K* studies included in the meta-analysis). The one-stage IPD can then be defined as:

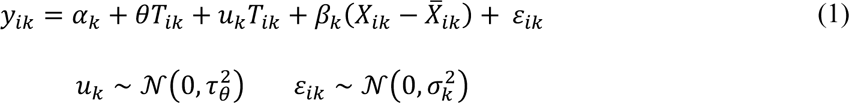

Where *T_ik_* is the treatment indicator, *X_ik_* is the symptom severity score at baseline, 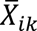 is the mean of *X* for each specific trial, α*_k_* is the stratified trial intercept, and *u_k_* is the trial-specific random slope of the average treatment effect θ. For this model, priors mirroring the two-stage approach will be used throughout, with a uniform distribution assigned to 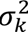.

The same model specification, but with a logit link, will be used to model the effects on binary outcomes, including remission, clinically meaningful improvement and clinically meaningful deterioration based on PHQ-9 thresholds:

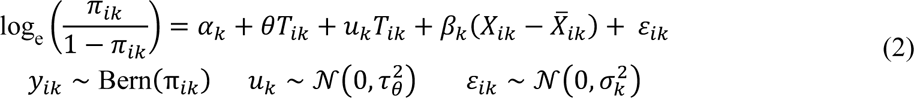

Where *y_ik_* now represents the patient’s binary outcome of interest. Numbers needed to treat (*NNT*) corresponding to the estimated SMD values will be calculated using the method by Furukawa and Leucht [62], and as the inverse of the absolute risk reduction otherwise.

### Moderator analyses

We will explore potential moderators of treatment effect by adding individual participant level and study level treatment-covariate interactions into the main IPD-MA model. To optimise power, these analyses will be done on treatment effects at post-treatment as all studies can provide data for this timepoint.

### Missing data

We will record the percentage of individual participant missing data for baseline characteristics and outcomes in each of the studies included in the meta-analyses. All analyses will be conducted according to the “intention-to-treat” principle, so that analyses target a treatment policy estimand [63]. Missing data will be handled using multiple imputation (fully conditional specification; MICE algorithm [64] under the missing at random (MAR) assumption. For the main effectiveness analysis, multilevel two-stage imputation models with heteroscedastic errors will be used to account for the nested data structure [65]. Highly collinear variables will be removed as predictors, as well as variables with systematically missing information (“structural zeros”). A total of *m*=50 imputation sets will be generated. A separate imputation model will be constructed for each focal moderator, including auxiliary variables with a maximum of 10% missingness. Imputations will be generated separately for each treatment groups (intervention, control) to account for treatment-covariate interactions. All analysis models will be fitted in the multiply imputed data. Parameter estimates and test statistics will then be aggregated by mixing the draws from the posterior distribution of each completed dataset [66]. Sensitivity analyses will be performed to compare results using observed and imputed data [67].

### Subgroup analyses

We plan to examine the efficacy of MBCT and potential moderators of treatment outcomes in subgroups defined by the conventional descriptive categories used by the individual studies to define their populations, distinguishing between studies that recruited patients characterised as treatment non-responders/remitters, treatment-resistant, or as suffering from chronic depression. These analyses will be based on subsamples created from the final IPD dataset.

### Sensitivity analyses

As a sensitivity analysis, we will repeat all main effectiveness analyses using equivalent frequentist (one- and two-stage) approaches. Additionally, we will conduct sensitivity analyses to investigate whether sources of heterogeneity affect the overall effect size estimate and robustness of our findings. This will include exploring the role of risk of bias and other potential study level sources of heterogeneity that may become obvious after data have been examined. These analyses will be conducted using a two-stage IPD-MA model.

## DISCUSSION

This IPD meta-analysis will provide estimates of efficacy for the use of MBCT in DTD and offer comprehensive information about potential moderators of treatment effects. As such it will contribute to arguments in favour of or against implementing MBCT as a treatment for DTD and may help to decide whether MBCT is a recommendable treatment option for this group. This is important given the need to improve the care and management of DTD and the increasing recognition that psychological therapies can have an integral role in this endeavour. Information about potential moderators of treatment effects will help to refine the understanding of who may or may not benefit from MBCT for DTD The chosen approach using IPD has several advantages over conventional meta-analyses including the possibility to assess whether individual participant level characteristics influence treatment effects with more specificity and power. Limitations of the approach include the fact that inconsistencies across studies regarding the format and type of variables reported may restrict our ability to investigate moderators of treatment effects and that transformation of outcome measures to a common metric across all studies may come with a small loss of information. To address this latter point, the current study will complement one-stage IPD analyses with a conventional two-stage approach. A more general concern that is often raised for IPD meta-analyses is inclusion bias as it may be difficult to obtain IPD for all selected studies.

To our knowledge, this is the first IPD meta-analysis to investigate the effects of MBCT in DTD and assess moderators of treatment effects. As evidence from trials of MBCT for DTD has now accumulated to a point where the use of MBCT for DTD can be considered for endorsement in treatment guidelines, findings from this IPD meta-analysis have the potential to inform treatment recommendations and decisions regarding implementation. The results will provide clinicians, healthcare providers and people who are suffering from DTD with information on the characteristics that may make it more or less likely for patients to benefit from this treatment and thus facilitate implementation.

## ETHICS AND DISSEMINATION

No local ethical review was necessary following consultation with the Ethics and Governance Board of the University of Surrey. The investigators of the primary trials have local ethical approval for sharing the data. TB and MH will be responsible for oversight of the database and analyses will be conducted by MH and TB. Patient privacy will be ensured by adhering to University of Surrey guidance on data management and storage. Exchange of data is governed by inter-institutional data sharing agreements. All data were pseudonymised (de-identified) before sharing, shared through secure upload facilities and are stored on secure servers. The data set will not be open access. Researchers of good standing may receive access pending on institutional approvals on data transfer.

We will disseminate the work in peer-reviewed journals, provide lay summaries and discuss findings with patient and lived experience groups. Other means of communication to disseminate the results will include conference presentations and social media announcements.

## CONTRIBUTIONS

TB, MN, JM, MV and MH conceptualised and designed the study. TB developed the search strategy and contacted the primary authors, who contributed to further refinement of the design and approach of the study. TB, MV and MH are responsible for building the database. MH provides statistical expertise and is responsible for data analyses. TB drafted the protocol manuscript with statistical input by MH, which was critically revised by all authors. All authors read and approved the final version. TB serves as the guarantor of the review.

## FUNDING STATEMENT

This research received no specific grant from any funding agency in the public, commercial or not-for-profit sectors.

## Data Availability

This is individual participant data from randomized controlled trials, which cannot be shared publicly due to ethical and consent restrictions. Interested parties will be asked to contact the corresponding author to request data access. Access may be granted if the following conditions are met:(1) all co-authors approve for the data sharing, (2) a data-sharing agreement is in place, (3) individual studies have participant consent and ethics approvals in place to allow for further onward sharing, and (4) an analysis plan has been developed and agreed upon by all co-authors. Once shared, the data may only be used for the specified purposes outlined in the agreement.

## ACKNOWLEDGMENTS

JMM has a ‘Miguel Servet’ research contract from the ISCIII (CP21/00080), and he is grateful to the Spanish CIBER of Epidemiology and Public Health (CBERESP CB22/02/00052< ISCIII) for their support.

## COMPETING INTERESTS STATEMENT

TB is the co-author of a book on mindfulness-based interventions and regularly offers workshops on mindfulness-based interventions. He is the co-investigator of a programme grant evaluating an adapted MBCT course for adolescents experiencing depression.

MN is author of a book chapter on mindfulness-based interventions and offers workshops on mindfulness-based interventions.

JM is the director of the Achtsamkeitsinstitut Ruhr (an institute offering mindfulness training) and principal investigator of several DFG (German Science Foundation) research projects. He receives royalties from mindfulness books he has authored.

MV reports no conflicts of interest.

JMGW co-developed MBCT, and was founding Director of the University of Oxford Mindfulness Centre. He receives royalties from books describing the MBCT program. He also receives honoraria from lectures and training workshops on mindfulness.

AC leads a professional practice (Istituto Mente Corpo) that offers courses of mindfulness-based interventions.

SE is the author of book on Mindfulness-Based Cognitive Therapy for the treatment of chronic and treatment-resistant depression.

KD declares no conflicts of interest.

ZS received book royalties from Guilford Press, workshop fees from the Centre for Mindfulness studies, and revenue from online sales at Mindful Noggin Inc., which are all related to his work as a co-founder of Mindfulness-Based Cognitive Therapy (MBCT).

MCM declares no conflicts of interest.

AS is the founder and director of the Radboudumc Expertise Centre for Mindfulness and co-investigator of a number of studies examining the effectiveness and implementation of mindfulness-based interventions for patients with psychological or somatic disorders and health care professionals.

AAF declares no conflicts of interest. MGT declares no conflicts of interest.

JMM is associated with the University of Oxford Mindfulness Research Centre. BDD is the lead of the University of Exeter AccEPT clinic, which offers courses of MBCT.

CS is a member of a training organisation that has been commissioned by NHS England to deliver MBCT training across NHS Talking Therapies services in England and co-leads the Sussex Mindfulness Centre. CS is a co-investigator of a programme grant evaluating an adapted MBCT course for adolescents experiencing depression and co-investigator on a grant evaluating an adapted MBCT course for NHS staff.

FAR regularly provides workshops on mindfulness-based interventions. MR declares no conflicts of interest.

MH serves as a statistical consultant for HelloBetter/GetOn Institut für Gesundheitstrainings GmbH, a company that implements digital mental health therapeutics in routine care.

---

**APPENDIX:** Search Terms.

Search Terms
| Database | Search Term |
| --- | --- |
| Web of Science | (“MDE” OR “major depressive episode” OR “MDD” OR “major depressive disorder” OR “major depression” OR “depression” OR “dysthymia” OR “dysthymic”) AND (“chronic” OR “persistent” OR “treatment-resistant” OR “non-responders” OR “non-response” OR “non-remitted” OR “dysthymia” OR “dysthymic”) AND (“MBCT” OR “mindfulness-based cognitive therapy”) AND (“randomized controlled trial” OR “randomised controlled trial” OR “randomized-controlled trial” OR “randomised-controlled trial” OR “RCT” OR “randomized” OR “randomised” OR “randomly allocated” OR “randomly assigned”) |
| Scopus | (“MDE” OR “major depressive episode” OR “MDD” OR “major depressive disorder” OR “major depression” OR “depression” OR “dysthymia” OR “dysthymic”) AND (“chronic” OR “persistent” OR “treatment-resistant” OR “non-responders” OR “non-response” OR “non-remitted” OR “dysthymia” OR “dysthymic”) AND (“MBCT” OR “mindfulness-based cognitive therapy”) AND (“randomized controlled trial” OR “randomised controlled trial” OR “randomized-controlled trial” OR “randomised-controlled trial” OR “RCT” OR “randomized” OR “randomised” OR “randomly allocated” OR “randomly assigned”) |
| PubMed | (“MDE” OR “major depressive episode” OR “MDD” OR “major depressive disorder” OR “major depression” OR “depression” OR “dysthymia” OR “dysthymic”) AND (“chronic” OR “persistent” OR “treatment-resistant” OR “non-responders” OR “non-response” OR “non-remitted” OR “dysthymia” OR “dysthymic”) AND (“MBCT” OR “mindfulness-based cognitive therapy”) AND (“randomized controlled trial” OR “randomised controlled trial” OR “randomized-controlled trial” OR “randomised-controlled trial” OR “RCT” OR “randomized” OR “randomised” OR “randomly allocated” OR “randomly assigned”) |
| PsycInfo | (“MDE” OR “major depressive episode” OR “MDD” OR “major depressive disorder” OR “major depression” OR “depression” OR “dysthymia” OR “dysthymic”) AND (“chronic” OR “persistent” OR “treatment-resistant” OR “non-responders” OR “non-response” OR “non-remitted” OR “dysthymia” OR “dysthymic”) AND (“MBCT” OR “mindfulness-based cognitive therapy”) AND (“randomized controlled trial” OR “randomised controlled trial” OR “randomized-controlled trial” OR “randomised-controlled trial” OR “RCT” OR “randomized” OR “randomised” OR “randomly allocated” OR “randomly assigned”) |
| Embase | (‘MDE’ OR ‘major depressive episode’ OR ‘MDD’ OR ‘major depressive disorder’ OR ‘major depression’ OR ‘depression’ OR ‘dysthymia’ OR ‘dysthymic’) AND (‘chronic’ OR ‘persistent’ OR ‘treatment-resistant’ OR ‘non-responders’ OR ‘non-response’ OR ‘non-remitted’ OR ‘dysthymia’ OR ‘dysthymic’) AND (‘MBCT’ OR ‘mindfulness-based cognitive therapy’) AND (‘randomized controlled trial’ OR ‘randomised controlled trial’ OR ‘randomized-controlled trial’ OR ‘randomised-controlled trial’ OR ‘RCT’ OR ‘randomized’ OR ‘randomised’ OR ‘randomly allocated’ OR ‘randomly assigned’) |
| Cochrane Controlled Trials | (“MDE” OR “major depressive episode” OR “MDD” OR “major depressive disorder” OR “major depression” OR “depression” OR “dysthymia” OR “dysthymic”) AND (“chronic” OR “persistent” OR “treatment-resistant” OR “non-responders” OR “non-response” OR “non-remitted” OR “dysthymia” OR “dysthymic”) AND (“MBCT” OR “mindfulness-based cognitive therapy”) AND (“randomized controlled trial” OR “randomised controlled trial” OR “randomized-controlled trial” OR “randomised-controlled trial” OR “RCT” OR “randomized” OR “randomised” OR “randomly allocated” OR “randomly assigned”) |

## Notes

### Author Declarations

No local ethical review was necessary following consultation with the Ethics and Governance Board of the University of Surrey.

